# Identification of multi-omic pleiotropy factors for peripheral artery disease

**DOI:** 10.1101/2025.07.11.25331391

**Authors:** Jiaqi Hu, Cassius I. Ochoa Chaar, Hongyu Zhao, Andrew T. DeWan

**Author notes:** To whom correspondence should be addressed: Dr. Andrew T. DeWan, Yale School of Public Health, Center for Perinatal, Pediatric and Environmental Epidemiology 1 Church Street, 6^208^ Floor, New Haven, Connecticut, 06510, USA.

## Abstract

**Background:** Peripheral artery disease (PAD) is prevalent and frequently co-occurs with type 2 diabetes (T2D) and coronary artery disease (CAD). Although shared genetic factors may contribute to these comorbidities, few studies have examined pleiotropy at the transcriptomic and proteomic levels.

**Methods:** We generated summary statistics for transcriptome-wide (TWAS) and proteome-wide (PWAS) association analyses across PAD, T2D, and CAD. Joint tests for pleiotropy at the variant, transcript, and protein levels were performed using PLACO, and the contributions of these pleiotropic factors to genetic correlations were quantified. Furthermore, we developed pleiotropy models integrating variants, transcripts, and proteins.

**Results:** In the PAD-T2D analysis, we identified 5 SNPs, 2 predicted transcripts, and 2 predicted proteins that together contributed 2.8% to the genome-wide genetic covariance. In the PAD-CAD analysis, 33 SNPs, 2 predicted transcripts, and 2 predicted proteins accounted for 9.15% of the genetic covariance. Overall, eight pleiotropy models were established, providing a multi-layered framework for categorizing our findings.

**Conclusion:** Employing a multi-omic framework, this study elucidates the shared genetic architecture of PAD with T2D and CAD. Our findings enhance the understanding of genetic risk factors underlying PAD and its comorbidities, with potential implications for future therapeutic strategies.

**Lay summary:** This study investigates why peripheral artery disease (PAD) often occurs together with type 2 diabetes (T2D) or coronary artery disease (CAD) by identifying shared genetic risk factors.

- We discovered new PAD-related genetic variants, including rs2138161, which may function through pathways shared between blood vessel function and metabolic processes.
- PAD-T2D comorbidity appears to be closely related to inflammatory pathways, while PAD-CAD comorbidity is more strongly linked to lipid-related pathways, offering insights into disease mechanisms and potential therapeutic targets.

**Graphical Abstract:** PAD: peripheral artery disease; T2D: type 2 diabetes; CAD: coronary artery disease; SNP: single nucleotide polymorphism; mRNA: messenger ribonucleic acid.

## Introduction

Peripheral artery disease (PAD) is a prevalent and severe disease, with the number of people living with PAD rising from 202 million in 2010^1^ to 236 million in 2015^2^. Type 2 diabetes (T2D) is a well-established risk factor for PAD^1,3^, with recent meta-analyses indicating a 1.96-fold increased risk of PAD in women and a 1.84-fold increase in men with diabetes^3^. Notably, up to 30% of PAD patients have diabetes^4^. In addition, PAD frequently co-occurs with coronary artery disease (CAD), with 39% of PAD patients diagnosed with CAD^5^. CAD is also a leading cause of death among PAD patients^5^. Understanding the comorbid relationship between T2D, PAD, and CAD may provide critical insights into the underlying etiology of PAD.

Pleiotropy, a phenomenon in which a single nucleotide polymorphism (SNP) or gene influences multiple diseases, may explain the observed comorbidities. The same individual SNPs and genes are related to both PAD and T2D^6–9^, as well as both PAD and CAD^8,10,11^. Polygenic scores (PGSs), which aggregate the effects of single genetic markers into an overall risk score, have been constructed for T2D and shown to be significantly associated with the PAD risk^6,9^. Similar findings have been reported for CAD PGSs^12^. The evidence supports a shared genetic risk for PAD-T2D and PAD-CAD comorbidities.

Pleiotropy falls into two main categories: vertical (also known as mediated) and horizontal (also known as biological). Vertical pleiotropy describes the scenario where the observed association between genetic factor and trait A is completely mediated by its association with trait B. Whereas horizontal pleiotropy describes the scenario where a genetic factor is independently associated with trait A and trait B, or rather, the genetic association with trait B is still observed even after adjusting for trait A. Horizontal pleiotropy can be further categorized into correlated and uncorrelated types, defined by whether a SNP or gene functions via the same molecular pathway in two diseases^9^.

Correlated pleiotropy involves SNPs functioning within a multi-omic network encompassing the genome, transcriptome, and proteome for both diseases. Uncorrelated pleiotropy consists of SNPs influencing independent pathways, for example, different transcripts and subsequent proteins. A comprehensive understanding of the molecular mechanisms underlying SNP-disease associations is crucial for revealing the complex etiology of PAD and its comorbidities. Identifying multi-omic pleiotropy factors across multi-omic levels can provide insights into convergent biological pathways that drive disease pathogenesis.

Various studies have been conducted to explore the associations or causal relationships across single- and multi-omic layers contributing to the risk of PAD, T2D, and CAD individually^13–18^. Despite these efforts, few studies have explored the pleiotropy between PAD-T2D and PAD-CAD at the transcriptomic and proteomic levels, making it challenging to categorize pleiotropic SNPs and genes into correlated or uncorrelated types or to model more complex relationships.

This study aims to address these gaps by exploring multi-omic risk profiles of PAD, T2D, and CAD through genome-, transcriptome- and proteome-wide association studies (TWAS and PWAS). We first establish disease-specific risk profiles and then identify and analyze pleiotropic genomic, transcriptomic, and proteomic factors. Our results not only enhance the understanding of PAD pathogenesis, but also elucidate the molecular mechanisms underlying its comorbidities—facilitating the identification of risk factors and therapeutic targets.

## Methods

### Overview of methods

Figure 1 presents the flowcharts for single-disease X-WAS and pleiotropy analyses. We conducted TWASs and PWASs for each single disease to generate summary statistics. Next, we examined pleiotropy for PAD-T2D and PAD-CAD comorbidities, assessing SNPs, predicted transcripts, and predicted proteins via PLACO. The factors identified were further evaluated for single-disease association and vertical pleiotropy. Validated pleiotropic components were quantified for their contributions to genome-wide genetic correlations and categorized into newly defined pleiotropy models.

**Figure 1.**
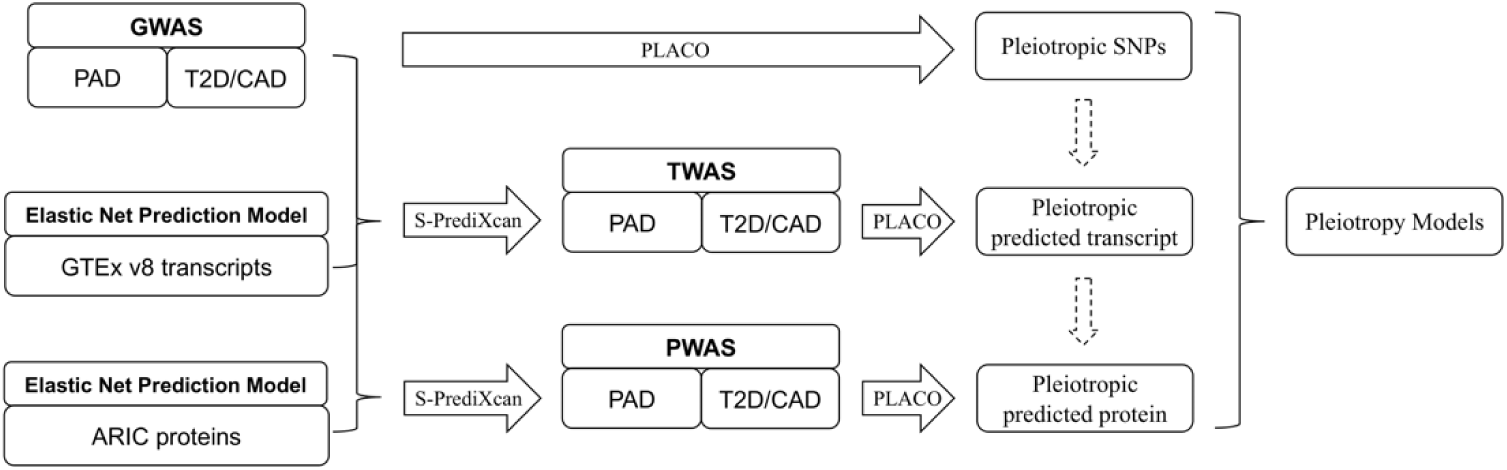
Flowchart of analysis. Flowchart of methodology. We first conducted TWAS and PWAS across three diseases, and ran PLACO on SNP, predicted transcripts, and predicted proteins for pleiotropy factors. Finally, we integrated pleiotropy results across multi-omic levels and developed pleiotropy models. GWAS: genome-wide association analysis; PAD: peripheral artery disease; T2D: type 2 diabetes; CAD: coronary artery disease; SNP: single-nucleotide polymorphism; TWAS: transcriptome-wide association analysis; PWAS: protein-wide association analysis.

### GWAS summary statistics

We conducted a meta-analysis of large-scale GWAS summary statistics from the Million Veteran Program (MVP)^8^ and UK Biobank (UKB) European studies on PAD using METAL^19^. The MVP is a large cohort study involving participants from 63 VA Medical Centers across the United States. Specifically, this GWAS includes 24,009 PAD cases and 150,983 non-PAD controls of Non-Hispanic White ancestry. Details of genotype quality control procedures are provided in the previous study^8^. After genotype imputation, SNPs were filtered according to strict quality criteria. A total of 17,577,362 SNPs were analyzed for associations with PAD using a logistic regression model adjusted for age, sex, and the top five genetic principal components (PCs). We accessed the summary statistics through dbGaP (dbGaP Study Accession: phs001672.v11, Approved Project Number: 32518).

The UKB study comprises genotype and phenotype data from 502,618 participants recruited from 22 assessment centers in the UK between 2006 and 2010^20^. Our analysis was restricted to subjects of White European ancestry, as defined by self-reported records and genetic ancestry described previously^21^. PAD cases and controls were identified using self-reports and medical records (the 9th and 10th versions of the International Classification of Diseases [ICD-9 and ICD-10], and OPCS-4) (Supplemental Table 1). We included 13,418,147 imputed SNPs with imputation scores ≥ 0.8 and Minor allele frequency (MAF) ≥ 0.001. We performed GWAS for PAD on the full set of directly genotyped and imputed variants (N=13,418,147) using REGENIE^22^. To adjust for confounding and potential bias due to population stratification, sex, age at recruitment, and the top 10 PCs calculated from the directly genotyped variants were included as covariates. The details can be found in the previously paper^23^.

For T2D and CAD, we used established large-scale GWAS summary statistics for European subjects^24,25^. Sample sizes for these analyses are provided in Table 1.

**Table 1.** GWAS summary statistics.

| Disease | GWAS | Sample sizes | Number of cases | Number of controls | Number of SNPs | Heritability (SE) | Genetic correlation with PAD (P-value) |
| --- | --- | --- | --- | --- | --- | --- | --- |
| PAD | Meta-analysis of MVP and UKB | 583,679 | 30,824 | 552,855 | 9,789,567 | 0.0153 (0.0041) |  |
| T2D | Mahajan et al. (2022) | 933,970 | 80,154 | 853,816 | 9,980,958 | 0.0419 (0.0032) | 0.5613 (2.17E-50) |
| CAD | Aragam et al. (2022) | 1,347,212 | 181,522 | 1,165,690 | 13,851,199 | 0.0321 (0.0027) | 0.6918 (4.54E-65) |

GWAS: genome-wide association study; SNP: single-nucleotide polymorphism; SE: standard error; PAD: peripheral artery disease; T2D: type 2 diabetes; CAD: coronary artery disease. The p-value for the genetic correlation is derived from a Wald test implemented within the LDSC framework.

### Single-omic X-WAS

We used the S-PrediXcan framework^26^ for TWAS and PWAS, integrating GWAS summary statistics and established prediction models from GTEx V8 whole blood tissue^27^ and Atherosclerosis Risk in Communities Study (ARIC) plasma proteins^28^, respectively. Details of the TWAS and PWAS can be found in the previous study^29^.

Lambda 1,000 (λ_1000_) was calculated for both TWAS and PWAS. λ_1000_ is a measurement of genomic inflation scaled to 1000 cases and 1000 controls using the equation: 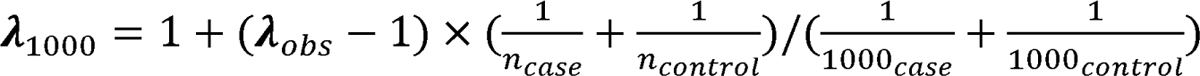 where λ_obs_ is the observed genomic inflation factor, and n_case_ and n_control_ are the number of cases and controls included in the GWAS summary statistics. A λ_1000_ less than or equal to 1.05 indicates an unbiased X-WAS^30^.

### Genome-wide genetic correlation

We calculated the genome-wide genetic correlations between PAD and CAD, as well as PAD and T2D, using LD Score regression (LDSC)^31^. The same GWAS summary statistics described earlier were used for this analysis, restricting to SNPs with MAFs greater than 0.05 in the summary statistics and those present in 1000 Genomes European genotype data. In total, 951,966 SNPs were included for the PAD-CAD correlation and 999,704 SNPs for the PAD-T2D correlation.

### Single-omic pleiotropy analysis

PLACO^32^ is a method that tests for pleiotropy by evaluating the null hypothesis that a variant is associated with at most one of two traits against the alternative hypothesis that it is associated with both traits. This method was used to identify pleiotropic SNPs with GWAS summary statistics for the diseases of interest. In brief, for each variant shared between two GWAS datasets, PLACO de-correlates the two Z scores and calculates an approximate asymptotic p-value for the hypothesis testing. Originally designed for SNP-level pleiotropy analysis, we extended PLACO to transcriptome and proteome levels using summary statistics from TWAS and PWAS. To account for multiple comparisons, we applied the FDR control method^33^ in an omic- and disease pair-specific manner.

PLACO uses the product of two Z scores for hypothesis testing, but one limitation of this method is that a strong association for one trait can disproportionately influence the result. In other words, if one SNP, predicted transcript, or predicted protein is strongly associated with one disease but not the other, PLACO may still generate a small p-value. To address this, we filtered out SNPs with p-values greater than 5×10^-6^ for either disease and excluded predicted transcripts or proteins with p-values greater than 0.05 for either disease. Additionally, we performed LD clumping via PLINK 1.9^34^ to obtain independent pleiotropic SNPs. We set the r^2^ threshold at 0.2 and used a default window size of 250 kilobases (kb). No p-value thresholds were set for index SNPs.

To investigate whether the pleiotropy signals identified were vertical or horizontal, we conducted adjusted association analyses using two models with individual-level genotype and phenotype data from UKB: (1) CAD/T2D ∼ PAD + SNP/predicted transcript/predicted protein + covariates, and (2) PAD ∼ CAD/T2D + SNP/predicted transcript/predicted protein + covariates. The covariates included age at recruitment, sex, and the top 10 genetic PCs. Due to our interest in dissecting signals between correlated and uncorrelated pleiotropy, we only focused on those signals that demonstrated some evidence of horizontal pleiotropy by only keeping those variants with at least nominally significant signals (p-values < 0.05) in both models.

### Contributions to genetic covariance

After validating the pleiotropic signals, we assessed their contributions to genome-wide genetic covariances using a leave-one-out approach. Specifically, we first identified SNPs in LD (r^2^>0.2) with the corresponding pleiotropic SNPs, eQTLs for pleiotropic predicted transcripts, or pQTLs for pleiotropic proteins. We then removed these SNPs from the GWAS summary statistics and calculated the genetic covariances using the truncated summary statistics through LDSC. We compared these leave-one-out genetic covariances with the genome-wide genetic covariances.

Furthermore, to determine whether the reduction in the number of SNPs could fully explain changes in genetic covariances, we conducted a permutation test with randomly selected regions. Specifically, we divided the summary statistics into near-independent LD regions through LD clumping and randomly selected the same number of regions as the pleiotropic components. SNPs within these regions were removed, and genetic covariances were recalculated from the summary statistics. The p-value for the permutation test was computed as the proportion of permutations in which the difference between the permuted genetic covariance and the overall genetic covariance exceeded the difference observed between pleiotropic genetic covariances and the overall genetic covariance. We conducted 1,000 permutations.

### Classification of pleiotropic signals

To better understand the molecular processes underlying pleiotropy, we classified pleiotropic components into different pleiotropy models. Each model consists of four layers: SNP, transcript, protein, and phenotype. In each layer, except for the phenotype layer, we assume either one or two signals. In total, we established eight pleiotropy models to reflect these different signal patterns (Figure 2).

**Figure 2.**
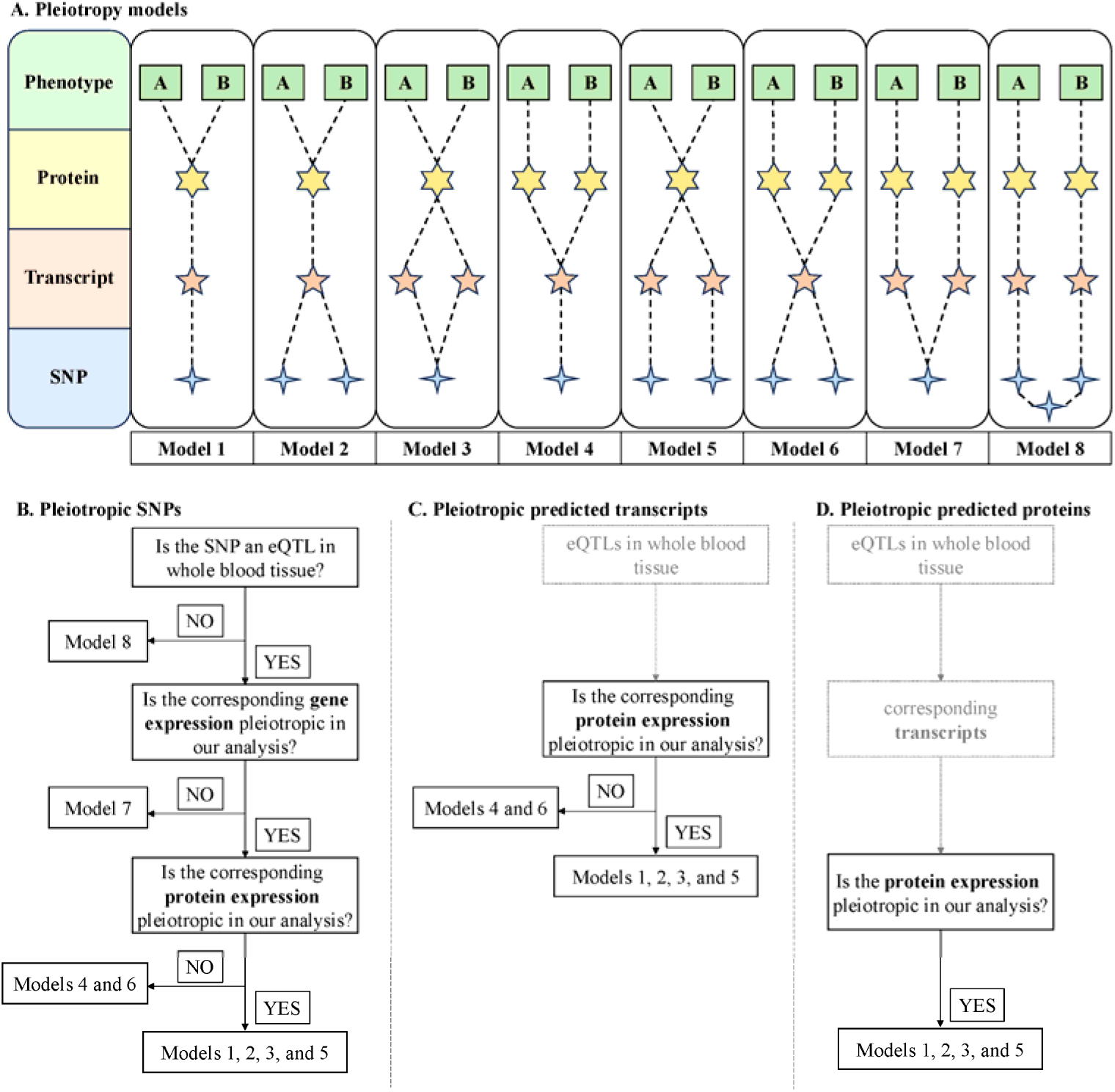
Pleiotropy models. Eight pleiotropy models integrating SNPs, transcripts, and proteins. (A). Definitions for eight pleiotropy models. Model 1 represents completely correlated pleiotropy, while Model 8 represents completely uncorrelated pleiotropy. Models 2 to 7 represent partially correlated pleiotropy, where one of two of the omic layers contains a single signal, while the remaining levels have two signals. (B-D). The assignment of specific factor to pleiotropy models.

Model 1 represents completely correlated pleiotropy, while Model 8 represents completely uncorrelated pleiotropy. Models 2 to 7 represent partially correlated pleiotropy, where one of two of the omic layers contains a single signal, while the remaining levels have two signals. We analyzed pleiotropic findings by omic level as follows.

We categorized pleiotropic SNPs into pleiotropic eQTLs and non-eQTLs based on GTEx v8 eQTLs in whole blood tissue. Pleiotropic eQTLs were classified into Models 1, 2, 3, and 5 if the corresponding gene(s) were pleiotropic in both TWAS and PWAS. They were assigned to Models 4 and 6 if the gene(s) were pleiotropic in TWAS but not in PWAS, and to Model 7 if not pleiotropic in either. Non-eQTLs were placed into Model 8, and we investigated potential eQTLs or pQTLs nearby (±500 kb) for additional details.

At the transcriptome level, we checked the PLACO results for the corresponding predicted proteins. Predicted transcripts were classified into Models 1, 2, 3, and 5 if the predicted protein(s) were pleiotropy. Otherwise, they were assigned to Models 4 and 6. For proteins, they were fitted to models 1, 2, 3, and 5. It is important to note that we assumed that the eQTLs for pleiotropic transcripts, pQTLs for pleiotropic proteins, and transcripts for pleiotropic proteins exhibit the same pleiotropic effects.

### Statistical analysis

Several sensitivity analyses were conducted to evaluate the robustness of our results. First, we conducted colocalization analyses for pleiotropic genes identified in TWAS and PWAS to assess consistency of findings across methods. Specifically, we analyzed whether eQTLs for genes identified in TWAS or pQTLs for genes identified in PWAS colocalized between the two diseases (PAD-CAD or PAD-T2D) using the coloc.abf function implemented in the coloc R package^35^. A posterior probability larger than 0.8 for either PP3 or PP4 suggests the presence of pleiotropy. Second, given that PLACO results may be affected by sample overlap between summary statistics, we applied PLACO+^36^, an enhanced version of PLACO designed to account for sample overlap, to re-evaluate our findings. Finally, we examined tissue specificity of the TWAS results by repeating analyses using TWAS models derived from disease-relevant tissues. For PAD, we used models from tibial artery tissue; for CAD, from aorta and coronary artery tissues; and for T2D, from pancreatic tissue. We then re-ran PLACO using genes initially identified in whole blood TWASs. Genes that remained significant across tissues were considered robust to the choice of tissue model.

To account for the increased type I error rate due to multiple comparisons, we applied the FDR control method^33^ unless otherwise specified. Most of the analyses were conducted using R version 4.2.

## Results

### Pleiotropy for PAD-T2D comorbidity

The same summary statistics were used to estimate the genetic correlations between PAD and T2D via LDSC. A strong positive correlation was found between PAD and T2D (0.56, p-value=2.17×10^-50^).

We generated TWAS and PWAS summary statistics that did not indicate any population stratification (1_1000_ < 1.05; Supplemental Figure 1). Next, we investigated specific SNPs, predicted transcripts, and predicted proteins that were pleiotropic for PAD and T2D. Among 8,407,719 shared SNPs between the GWAS summary statistics for T2D and PAD, 11,924 exhibited significant pleiotropic effects as identified by PLACO (p-value ≤ 7.09×10^-5^), and 711 SNPs had univariate GWAS p-values less than 5×10^-6^ for both PAD and T2D. After LD clumping, 15 independent pleiotropic SNPs remained. Applying PLACO to TWAS summary statistics identified 27 pleiotropic predicted transcripts (p-value ≤ 1.75×10^-4^), of which 25 were nominally significant in single-disease TWAS analyses. PLACO also revealed 6 pleiotropic predicted proteins, all significantly associated with both PAD and T2D (p-value < 0.05). We further validated the pleiotropic components using two regression models. We kept 5 SNPs, 2 predicted transcripts, and 2 predicted proteins that were nominally significant in both models (Table 2). We further conducted several sensitivity analyses to assess the robustness of our results. In the colocalization analysis, three genes (*C4A*, *DDAH2* and *APOE*) showed high posterior probabilities (>0.8) for either hypothesis 3 or 4, suggesting evidence of pleiotropy signals (Supplemental Table 2). In contrast, *PLEKHA1* exhibited a high posterior probability for hypothesis 2 (0.96), suggesting that additional validation is needed to confirm its pleiotropic role. When repeating the analysis using PLACO+, all the variants and genes we identified remained significant (<0.05), supporting the robustness of our findings to potential sample overlaps (Supplemental Table 3). When assessing the tissue specificity, *C4A* remained significant (p-value=4.4×10^-6^) when using TWAS models derived from tibial artery tissue for PAD and pancreatic tissue for T2D. In contrast, *DDAH2* did not have a valid prediction model available in either tissue, precluding further evaluation.

**Table 2.** Pleiotropy findings.

| Chr | Pos | SNP | A1/pos_end* | z.pad | p.pad | z.D2 | p.D2 | p.placo | D2 | study | Pleiotropy Model(s) |
| --- | --- | --- | --- | --- | --- | --- | --- | --- | --- | --- | --- |
| 2 | 226230443 | rs2138161 | C | 4.86 | 1.20E-06 | 13.77 | 3.72E-43 | 6.47E-21 | T2D | GWAS | 8 |
| 9 | 22119196 | rs1333045 | C | 12.12 | 7.84E-34 | 7.92 | 2.34E-15 | 2.65E-31 | T2D | GWAS | 8 |
| 9 | 133270061 | rs582118 | A | -5.06 | 4.09E-07 | -6.88 | 5.96E-12 | 2.60E-12 | T2D | GWAS | 7 |
| 11 | 48467808 | rs149027146 | T | 4.68 | 2.82E-06 | 4.77 | 1.83E-06 | 2.25E-08 | T2D | GWAS | 8 |
| 17 | 67964738 | rs59956089 | C | 6.74 | 1.61E-11 | 6.29 | 3.23E-10 | 9.34E-15 | T2D | GWAS | 8 |
| 6 | 31727038 | DDAH2 | 31730617 | -3.46 | 5.41E-04 | -6.44 | 1.21E-10 | 3.03E-06 | T2D | TWAS | 1-6 |
| 6 | 31982057 | C4A | 32002681 | -4.65 | 3.26E-06 | -5.79 | 7.08E-09 | 1.43E-07 | T2D | TWAS | 1-6 |
| 10 | 122374696 | PLEKHA1 | 122432354 | -3.09 | 2.00E-03 | 4.00 | 6.35E-05 | 1.96E-05 | T2D | PWAS | 1, 2, 3, 5 |
| 19 | 44905791 | APOE | 44909393 | 2.61 | 9.13E-03 | -4.71 | 2.51E-06 | 1.71E-05 | T2D | PWAS | 1, 2, 3, 5 |
| 1 | 150567391 | rs932054 | G | 4.65 | 3.34E-06 | 6.71 | 1.89E-11 | 3.71E-11 | CAD | GWAS | 7 |
| 2 | 226230443 | rs2138161 | C | 4.86 | 1.20E-06 | 7.11 | 1.17E-12 | 3.39E-12 | CAD | GWAS | 8 |
| 4 | 147480038 | rs6841581 | A | 6.26 | 3.86E-10 | 11.44 | 2.65E-30 | 2.18E-23 | CAD | GWAS | 8 |
| 6 | 34809756 | rs9296133 | A | 5.00 | 5.67E-07 | 6.32 | 2.64E-10 | 2.34E-11 | CAD | GWAS | 7 |
| 6 | 160003858 | rs144078421 | A | 6.10 | 1.04E-09 | 13.23 | 5.80E-40 | 1.31E-25 | CAD | GWAS | 7 |
| 6 | 160130061 | rs4646272 | G | 5.47 | 4.42E-08 | 10.61 | 2.55E-26 | 4.11E-19 | CAD | GWAS | 7 |
| 6 | 160298560 | rs79390162 | T | 7.61 | 2.78E-14 | 13.05 | 6.16E-39 | 4.51E-32 | CAD | GWAS | 8 |
| 6 | 160330499 | rs9295128 | T | 8.89 | 6.08E-19 | 21.96 | 6.60E-107 | 2.42E-58 | CAD | GWAS | 7 |
| 6 | 160338808 | rs386453 | T | -5.22 | 1.83E-07 | -10.63 | 2.19E-26 | 3.46E-18 | CAD | GWAS | 8 |
| 6 | 160446240 | rs143665477 | C | 5.23 | 1.67E-07 | 9.37 | 7.55E-21 | 1.73E-16 | CAD | GWAS | 8 |
| 6 | 160470865 | rs182443492 | A | 4.80 | 1.55E-06 | 10.18 | 2.48E-24 | 4.05E-16 | CAD | GWAS | 8 |
| 6 | 160589086 | rs10455872 | G | 14.51 | 9.74E-48 | 29.12 | 2.18E-186 | 1.17E-128 | CAD | GWAS | 8 |
| 6 | 160591981 | rs140570886 | C | 10.79 | 3.67E-27 | 22.45 | 1.36E-111 | 3.88E-74 | CAD | GWAS | 7 |
| 6 | 160646860 | rs6905073 | T | -4.74 | 2.17E-06 | -11.14 | 8.40E-29 | 5.32E-17 | CAD | GWAS | 8 |
| 6 | 160741258 | rs1835346 | G | 4.78 | 1.79E-06 | 6.79 | 1.14E-11 | 1.50E-11 | CAD | GWAS | 8 |
| 6 | 160962047 | rs143843429 | G | 6.70 | 2.09E-11 | 15.20 | 3.79E-52 | 1.17E-31 | CAD | GWAS | 8 |
| 7 | 19009765 | rs2107595 | A | 6.59 | 4.27E-11 | 10.14 | 3.61E-24 | 2.71E-22 | CAD | GWAS | 8 |
| 7 | 150993088 | rs3918226 | T | 5.15 | 2.56E-07 | 11.79 | 4.60E-32 | 1.88E-19 | CAD | GWAS | 8 |
| 9 | 21961867 | rs7041637 | A | 6.75 | 1.46E-11 | 18.01 | 1.52E-72 | 2.24E-36 | CAD | GWAS | 8 |
| 9 | 22069145 | rs16905599 | A | -4.65 | 3.28E-06 | -7.70 | 1.32E-14 | 1.68E-12 | CAD | GWAS | 8 |
| 9 | 22098575 | rs4977574 | G | 12.96 | 2.00E-38 | 35.60 | 1.41E-277 | 1.91E-132 | CAD | GWAS | 8 |
| 10 | 98130980 | rs11189490 | A | -5.70 | 1.18E-08 | -5.84 | 5.36E-09 | 6.05E-12 | CAD | GWAS | 7 |
| 11 | 102924877 | rs1892971 | A | -5.21 | 1.87E-07 | -6.21 | 5.14E-10 | 1.25E-11 | CAD | GWAS | 8 |
| 12 | 111466567 | rs4766578 | A | -6.94 | 3.85E-12 | -11.06 | 2.03E-28 | 2.66E-25 | CAD | GWAS | 8 |
| 12 | 112049014 | rs17696736 | G | 6.56 | 5.37E-11 | 9.30 | 1.36E-20 | 1.39E-20 | CAD | GWAS | 8 |
| 15 | 78643826 | rs55988292 | G | 5.40 | 6.48E-08 | -4.96 | 6.95E-07 | 5.86E-13 | CAD | GWAS | 7 |
| 16 | 56959249 | rs12149545 | A | -4.71 | 2.47E-06 | -6.70 | 2.11E-11 | 2.83E-11 | CAD | GWAS | 8 |
| 18 | 22422981 | rs1893250 | C | 5.22 | 1.75E-07 | 5.83 | 5.54E-09 | 5.01E-11 | CAD | GWAS | 8 |
| 18 | 22476030 | rs2110135 | C | -5.43 | 5.58E-08 | -5.88 | 4.14E-09 | 1.66E-11 | CAD | GWAS | 8 |
| 19 | 10959957 | rs111516643 | T | -4.57 | 4.92E-06 | -5.50 | 3.89E-08 | 2.79E-09 | CAD | GWAS | 8 |
| 19 | 11073161 | rs73015007 | A | -5.84 | 5.14E-09 | -7.95 | 1.82E-15 | 4.89E-16 | CAD | GWAS | 1-7 |
| 19 | 11077441 | rs55997232 | T | -6.23 | 4.62E-10 | -13.97 | 2.33E-44 | 2.40E-27 | CAD | GWAS | 8 |
| 19 | 11166556 | rs4804573 | A | -4.65 | 3.32E-06 | -9.00 | 2.18E-19 | 3.74E-14 | CAD | GWAS | 8 |
| 5 | 17107360 | CTD-2228A4.1 | 17107789 | -2.53 | 1.15E-02 | 4.44 | 8.83E-06 | 1.67E-05 | CAD | TWAS | 1-6 |
| 8 | 19901717 | LPL | 19967259 | -4.08 | 4.50E-05 | -7.29 | 3.01E-13 | 1.32E-07 | CAD | TWAS | 1-6 |
| 10 | 122374696 | PLEKHA1 | 122432354 | -3.09 | 2.00E-03 | -5.11 | 3.20E-07 | 1.65E-04 | CAD | PWAS | 1, 2, 3, 5 |
| 17 | 42313324 | STAT3 | 42388568 | -3.61 | 3.01E-04 | -4.22 | 2.49E-05 | 1.70E-04 | CAD | PWAS | 1, 2, 3, 5 |

Chr: chromosome; Pos: position in hg38; \*: effective allele for SNPs and ending position for genes. P-values for PAD, T2D, and CAD were obtained using Wald tests. The p-value for PLACO was derived from a likelihood-based composite test statistic.

We assessed the contributions of 9 pleiotropic components to the genome-wide genetic covariance between PAD and T2D using a leave-one-out strategy. These components in aggregate accounted for 2.8% of the genetic covariance, with a significant p-value from the permutation test (p-value < 0.001) (Table 3). Notably, rs1333045 and rs2138161 each explained 1.4%, ranking highest among all components. These results further validated the pleiotropic effects of the identified components.

**Table 3.** Contributions to genetic covariance.

| Disease | Genetic covariance | SNP/Gene | Number of SNPs removed | Genetic covariance after removing SNPs | Number of SNPs removed in permutation | Permuted genetic covariance | P-value |
| --- | --- | --- | --- | --- | --- | --- | --- |
| CAD | 0.0153 | All | 903 | 0.0139 | 457.34 | 0.0153 | 0 |
|  |  | rs4977574 | 20 | 0.0148 | 6.11 | 0.0153 | 0 |
|  |  | LPL | 102 | 0.0152 | 48.06 | 0.0153 | 0.002 |
|  |  | STAT3 | 197 | 0.0152 | 133.92 | 0.0153 | 0.006 |
|  |  | rs143665477 | 46 | 0.0152 | 6.12 | 0.0153 | 0 |
|  |  | rs386453 | 53 | 0.0152 | 6.12 | 0.0153 | 0 |
|  |  | rs9295128 | 46 | 0.0152 | 6.12 | 0.0153 | 0 |
|  |  | CTD-2228A4.1 | 17 | 0.0153 | 6.13 | 0.0153 | 0 |
|  |  | PLEKHA1 | 115 | 0.0153 | 69.38 | 0.0153 | 0.004 |
|  |  | rs10455872 | 17 | 0.0153 | 6.12 | 0.0153 | 0 |
|  |  | rs111516643 | 26 | 0.0153 | 5.98 | 0.0153 | 0.001 |
|  |  | rs11189490 | 22 | 0.0153 | 6.13 | 0.0153 | 0 |
|  |  | rs12149545 | 3 | 0.0153 | 5.97 | 0.0153 | 0.001 |
|  |  | rs140570886 | 17 | 0.0153 | 6.12 | 0.0153 | 0 |
|  |  | rs143843429 | 11 | 0.0153 | 6.12 | 0.0153 | 0 |
|  |  | rs144078421 | 9 | 0.0153 | 6.12 | 0.0153 | 0 |
|  |  | rs17696736 | 2 | 0.0153 | 6.05 | 0.0153 | 0 |
|  |  | rs182443492 | 9 | 0.0153 | 6.12 | 0.0153 | 0 |
|  |  | rs1835346 | 14 | 0.0153 | 6.12 | 0.0153 | 0 |
|  |  | rs1892971 | 1 | 0.0153 | 6.11 | 0.0153 | 0 |
|  |  | rs1893250 | 1 | 0.0153 | 5.94 | 0.0153 | 0.001 |
|  |  | rs2107595 | 12 | 0.0153 | 6.14 | 0.0153 | 0 |
|  |  | rs2110135 | 13 | 0.0153 | 5.94 | 0.0153 | 0.001 |
|  |  | rs2138161 | 48 | 0.0153 | 6.11 | 0.0153 | 0 |
|  |  | rs3918226 | 9 | 0.0153 | 6.14 | 0.0153 | 0 |
|  |  | rs4646272 | 0 | 0.0153 | 6.12 | 0.0153 | 0 |
|  |  | rs4766578 | 3 | 0.0153 | 6.05 | 0.0153 | 0 |
|  |  | rs4804573 | 3 | 0.0153 | 5.98 | 0.0153 | 0.001 |
|  |  | rs55988292 | 8 | 0.0153 | 6.00 | 0.0153 | 0.001 |
|  |  | rs55997232 | 4 | 0.0153 | 5.98 | 0.0153 | 0.001 |
|  |  | rs6841581 | 4 | 0.0153 | 6.10 | 0.0153 | 0 |
|  |  | rs6905073 | 17 | 0.0153 | 6.12 | 0.0153 | 0 |
|  |  | rs7041637 | 6 | 0.0153 | 6.11 | 0.0153 | 0 |
|  |  | rs73015007 | 4 | 0.0153 | 5.98 | 0.0153 | 0.001 |
|  |  | rs79390162 | 23 | 0.0153 | 6.12 | 0.0153 | 0 |
|  |  | rs9296133 | 45 | 0.0153 | 6.14 | 0.0153 | 0 |
|  |  | rs932054 | 21 | 0.0153 | 6.06 | 0.0153 | 0 |
|  |  | rs16905599 | 7 | 0.0154 | 6.11 | 0.0153 | 0 |
| T2D | 0.0143 | All | 508 | 0.0139 | 324.15 | 0.0143 | 0 |
|  |  | rs1333045 | 21 | 0.0141 | 5.83 | 0.0143 | 0 |
|  |  | rs2138161 | 51 | 0.0141 | 6.21 | 0.0143 | 0 |
|  |  | C4A | 216 | 0.0142 | 207.66 | 0.0143 | 0.001 |
|  |  | APOE | 11 | 0.0143 | 13.25 | 0.0143 | 0.002 |
|  |  | DDAH2 | 28 | 0.0143 | 34.95 | 0.0143 | 0.002 |
|  |  | PLEKHA1 | 119 | 0.0143 | 63.33 | 0.0143 | 0.001 |
|  |  | rs149027146 | 42 | 0.0143 | 5.90 | 0.0143 | 0 |
|  |  | rs582118 | 25 | 0.0143 | 5.87 | 0.0143 | 0 |
|  |  | rs59956089 | 16 | 0.0143 | 6.16 | 0.0143 | 0 |

P-values were obtained using a permutation test based on 1,000 iterations.

We defined 8 pleiotropy models (Figure 2 A) and classified the pleiotropic components across omic layers (Figure 2 B-D). Among the 5 pleiotropic SNPs, rs582118 was an eQTL for *ABO* in the whole blood tissue; however, the predicted expression of *ABO* was not associated with either PAD or T2D, placing it with Model 7. The remaining 4 SNPs were not eQTLs in the whole blood tissue and were categorized under Model 8. Notably, rs149027146 was in LD with eQTLs for *ACP2*, *C1QTNF4*, *MYBPC3*, *NUP160*, *PTPRJ*, and *SLC39A13*. The predicted expressions of the latter five genes were nominally associated with T2D, but none were associated with PAD, indicating the need to explore additional eQTLs related to PAD near this variant. For pleiotropic transcripts, the corresponding proteins for both *DDAH2* and *C4A* were not mapped in ARIC. No specific model among Models 1 to 6 can be assigned to these two genes. For pleiotropic protein expressions, *APOE* and *PLEKHA1* were classified under Models 1, 2, 3, and 5.

### Pleiotropy for PAD-CAD comorbidity

The genome-wide genetic correlation between PAD and CAD was 0.69 (p-value=4.54×10^-65^). The summary statistics of GWAS, TWAS, and PWAS for CAD also showed stable 1_1000_ (Supplemental Figure 1). We then applied PLACO to the summary statistics for PAD and CAD. After controlling the FDR at 0.05, we identified 17,746 pleiotropic SNPs, 43 pleiotropic predicted transcripts, and 9 pleiotropic predicted proteins. To refine the results, we excluded SNPs with p-values ≥ 5×10^-6^ and predicted transcripts or proteins with p-values ≥ 0.05 for both PAD and CAD. We then performed LD clumping to remove correlated SNP. In total, 60 SNPs, 40 predicted transcripts, and 9 predicted proteins were kept for further analyses. In addition, we explored the potential for vertical pleiotropy by assessing associations with one disease while adjusting for the other. Components with at least nominal significance (p < 0.05) in both models were retained. The final list consisted of 33 SNPs, 2 predicted transcripts, and 2 predicted proteins (Table 2). Among the four identified genes, *LPL* was the only one showing clear evidence of pleiotropy in the colocalization analyses (Supplemental Table 2). *CTD-2228A4.1* and *STAT3* exhibited modest pleiotropic signals, with the highest posterior probabilities corresponding to hypothesis 4, whereas *PLEKHA1* demonstrated a higher probability for hypothesis 2. Furthermore, our findings were robust to potential sample overlaps, as all associations remained significant when re-evaluated using PLACO+ (Supplemental Table 3). Neither *CTD-2228A4.1* nor *LPL* had valid prediction models available in the disease-relevant tissues and therefore no tissue-specificity analysis was conducted.

The leave-one-out analysis revealed that, collectively, 37 components explained 9.15% of the genetic covariance between PAD and CAD (permutation p-value<0.001) (Table 3). The top contributing component was rs4977574, accounting for 3.27% (permutation p-value = 0). Notably, rs16905599 had a negative contribution to the genetic covariance (−0.65%, permutation p-value < 0.001). These results further supported the pleiotropy of identified components for PAD-CAD.

Among the 33 pleiotropic SNPs, 9 were identified as eQTLs in whole blood tissue, with 4 of these associated with the expression of *SLC22A1*. These corresponding genes were not significant in the PLACO analysis, suggesting Model 7, except for one SNP, rs111516643, which was an eQTL for *SMARCA4*–a gene not analyzed in TWAS. The current results were not sufficient to distinguish between Models 1 to 7 for rs111516643.

The remaining 24 SNPs were not eQTLs in whole blood tissue and were assigned to Model 8. Specifically, rs17696736 was in LD with eQTLs for *ALDH2*, *MAPKAPK5-AAS1*, and *TMEM116*, but none were significantly associated with PAD in the TWAS, suggesting other nearby genes linked to PAD. Regarding the two pleiotropic predicted transcripts, *CTD-2228a4.1* was not a protein-coding gene and might regulate the protein expression of other genes through Models 1 to 6. The protein for *LPL* was not mapped in ARIC, and we cannot distinguish Models 1 to 6. The pleiotropic predicted protein expressions of *PLEKHA1* and *STAT3* were assigned to Models 1, 2, 3, and 5.

## Discussion

This study employed a multi-omic framework to investigate the shared genetic architecture between PAD and two related diseases–T2D and CAD. For PAD-T2D, we identified 5 SNPs, 2 predicted transcripts, and 2 predicted proteins that collectively contributed 2.8% to the genome-wide genetic covariance. Similarly, the PAD-CAD analysis revealed 33 SNPs, 2 predicted transcripts, and 2 predicted proteins, accounting for 9.15% of the genetic covariance. Furthermore, we proposed eight pleiotropy models and categorized our findings accordingly. These results enhance our understanding of the genetic risk profiles of PAD with its comorbidities, T2D and CAD.

Analyzing PAD alongside related diseases enhances the power to identify risk factors and facilitates the discovery of variants and genes not previously linked to PAD. For example, our analysis revealed that rs2138161 was associated with both PAD and T2D despite limited prior evidence of its involvement in PAD. Notably, previous studies have demonstrated that the C allele of rs2138161 is linked to an increased risk of metabolic syndrome among European subjects in UKB^37^, an increased risk of cardiometabolic multimorbidity (CMM), defined as the coexistence of more than two diseases of T2D, CAD, and stroke, in UKB subjects^38^, and elevated levels of multiple lipid metabolites^39^. These findings suggest that rs2138161 may play a role in shared biological pathways underlying metabolic and vascular dysfunction. This observation underscores the value of pleiotropic analyses in uncovering genetic risk factors for complex diseases and highlights the need for further functional studies to elucidate the mechanistic role of rs2138161 in PAD pathogenesis.

Further, the predicted protein expression level of *PLEKHA1* was positively associated with both PAD and CAD but inversely associated with T2D. This gene has been previously implicated in T2D^40^, osteoporosis^40^, and neurodegenerative disorders such as Alzheimer’s disease (AD) and age-related macular degeneration^41^. *PLEKHA1* is closely related to the PI3K/AKT signaling pathway–a key mediator in the development of ischemic diseases affecting the heart, brain, and limbs and a recognized therapeutic target^42,43^. Our results suggest that the PLEKHA1 protein may serve as a potential therapeutic target for PAD via modulation of the PI3K/AKT pathway. This differential association across related conditions underscores the complex role of *PLEKHA1* in both vascular and metabolic pathogenesis. It highlights the need for further mechanistic studies to clarify its contributions to these disease processes.

Our study identified genetic components underlying the comorbidity of PAD with T2D and CAD. In the analysis for PAD and T2D, we have identified two predicted transcripts (*C4A* and *DDAH2*) and two predicted proteins (*APOE* and *PLEKHA1*) as pleiotropy factors, with all four genes previously linked to AD^41,44–47^. Our findings are consistent with earlier research demonstrating overlapping genetic risk among cardiovascular diseases, AD, and T2D^41,48^, which supports the idea of common pathogenic mechanisms. Chronic inflammation–a known contributor to AD, atherosclerosis, and T2D^49,50^–may serve as a central mediator of this genetic overlap. Future studies of the effects of similar inflammatory pathways on the comorbidity of PAD and T2D need to be conducted to clarify the complex mechanisms further.

Lipids and lipoproteins play important roles in the pathogenesis of atherosclerosis^51,52^. Our analyses identified several lipid-related genetic factors that exhibit pleiotropy effects on PAD and CAD. For example, we found rs140570886, a well-established risk variant for elevated lipoprotein (a) levels located in the *LPA* gene region^53^, that was associated with an increased risk of both PAD and CAD. In addition, the predicted transcript level of *LPL* was inversely associated with these two diseases. This gene encodes lipoprotein lipase, a key enzyme in lipid metabolism that also influences atherosclerotic development^54,55^. Our results suggest that pathways involved in lipid metabolism may underlie the shared genetic basis of PAD and CAD, underscoring the clinical importance of effective lipid management in atherosclerotic patients.

The molecular basis of pleiotropy has been extensively discussed. Traditional models categorize a mutation’s impact on multiple phenotypes into three tiers–gene, gene product, and function^56^. In our study, we extended this framework by adopting a multi-omic perspective that integrates SNPs, transcripts, and proteins (Figure 2). Our multi-layered models clarify how genetic variation concurrently influences two complex diseases through changes in gene and protein expression. By elucidating these molecular mechanisms, our framework highlights potential biological pathways and therapeutic targets for future research.

A major strength of our study is the application of joint tests using PLACO to assess whether genetic variants, predicted transcripts, or predicted proteins are simultaneously associated with two diseases. Prior investigations into PAD-related pleiotropy have predominantly relied on candidate SNP association^10^, MR^9^, multi-trait association^11^, or PGS analyses^9,12^. In contrast, few studies have employed joint tests specifically designed to detect pleiotropy at multiple molecular levels. Our findings provide direct statistical evidence of pleiotropy, and we extend the PLACO framework to encompass transcriptomic and proteomic analyses. This multi-omic approach not only reinforces the evidence of shared genetic underpinnings but also offers new insights into the molecular mechanisms driving the comorbidities of PAD with T2D and CAD.

Several limitations should be acknowledged. First, the TWAS and PWAS summary statistics are derived from models that predict gene and protein expression levels based on genetic variants rather than from direct measurements. As such, the accuracy of these predictions is critical. Additionally, these predicted expression levels should not be interpreted as direct measurements due to variations in the heritability of gene and protein expression and the exclusion of non-genetic risk factors from the models. Instead, the observed associations reflect the relationship between the genetic component of omic levels and disease risk.

Second, our pleiotropy models integrate each component into a single framework, implicitly assuming that these components act independently. Additionally, we focused exclusively on cis effects for both transcripts and proteins in our prediction and subsequent analyses, despite the potential significance of trans effects in mediating pleiotropy. Future studies should incorporate gene interactions and trans effects to capture a more comprehensive picture of pleiotropic mechanisms.

Third, we did not perform simulation analyses to evaluate the false positive rate or statistical power of extending PLACO to TWAS and PWAS, as such evaluations were beyond the scope of this study. To assess the robustness of the pleiotropic gene and protein associations identified by our method, we conducted colocalization analyses. Most results showed at least suggestive evidence of pleiotropy, whereas the predicted protein expression of *PLEKHA1* did not demonstrate evidence of pleiotropy in either the PAD-T2D or PAD-CAD pairs. Further studies should be conducted to more comprehensively evaluate the performance of PLACO in multi-omic contexts.

Fourth, gene and protein expression levels vary across tissue types, and therefore our findings based on whole-blood TWAS and PWAS may not fully generalize to other disease-relevant tissues. To evaluate potential tissue specificity, we conducted a sensitivity analysis using TWAS models derived from tissues pathologically relevant to each disease. Among the four genes identified, only *C4A* had a valid prediction model in each of the selected tissues, and its pleiotropic association was successfully replicated. The remaining three genes, although expressed in these tissues, lacked valid prediction models and thus could not be evaluated. Future studies incorporating more comprehensive and tissue-specific transcriptomic resources will be important for further investigating the tissue specificity of these pleiotropy signals.

Lastly, our study was limited to genetically confirmed European subjects due to the small sample sizes of GWAS in other populations and the absence of gene and protein expression prediction models for non-European groups. Given the differences in disease prevalence, heritability, and genetic architecture across ancestries, our findings may not be generalizable to non-European populations. Future studies will aim to incorporate larger, more diverse datasets to validate and extend these results.

## Conclusions

To sum up, our multi-omic analysis investigated pleiotropic genetic factors associated with two disease pairs–PAD with T2D and PAD with CAD. By integrating the results of joint tests across SNP, predicted transcript, and predicted protein levels, we developed eight pleiotropy models to elucidate the molecular process involved in pleiotropy. These findings provide valuable insights into the genetic etiology of both individual diseases and their comorbidities.

## Data Availability

The GWAS summary statistic used in this study is available through the CARDIoGRAMplusC4D website (http://www.cardiogramplusc4d.org/) for CAD and the DIAGRAM website (https://diagram-consortium.org/downloads.html) for T2D. The GWAS summary statistic for PAD developed using the MVP cohort is available via application through dbGaP (dbGaP Study Accession: phs001672.v11). The meta-analyzed GWAS for PAD used in this study is available from the corresponding author upon reasonable request. The elastic net models used in TWAS can be download via https://zenodo.org/records/3519321#.XfEKwtF7m90. The PWAS prediction models can be downloaded via http://nilanjanchatterjeelab.org/pwas/. The TWAS and PWAS summary statistics generated in this study are available from the corresponding author upon reasonable request.

## Acknowledgements

The authors would like to thank the researchers and participants of the United Kingdom Biobank. All data was accessed as part of project 32285 from the United Kingdom Biobank. We thank the Yale Center for Research Computing for the use of the McCleary High Performance Computing cluster.

## Funding Statement

This work was supported by a grant from the National Institutes of Health-National Heart, Lung, and Blood Institute (R01HL145660 to ATD), a grant from National Institutes of Health-National Institute of General Medical Sciences (R01GM134005 to HZ), and a grant from National Institutes of Health-National Human Genome Research Institute (R01HG012735 to HZ).

## Author Contributions

Conceptualization: JH, HZ, AD; Data curation: JH, AD; Formal analysis: JH; Funding acquisition: HZ, AD; Investigation: JH, HZ, AD; Methodology: JH, HZ, AD; Project administration: AD; Resources: HZ, AD; Software: JH; Supervision: HZ, AD; Validation: JH; Visualization: JH, AD; Wring-original draft: JH, AD; Wriging-review & editing: all authors.

## Ethics Declaration

This research was conducted using the UK Biobank Resource (application number 32285). The UK Biobank study was conducted under generic approval from the National Health Services’ National Research Ethics Service. The present analyses were conducted in accordance with the Declaration of Helsinki and approved by the Human Investigations Committee at Yale University (2000026836 for UK Biobank). The patients/participants provided their written informed consent to participate in this study.

## Conflicts of interest

CIOC is consultant for SVS-PSO, EnVVeno Medical, has IP of patent U.S.S.N. 10,524,89, and has received research support from Yale department of Surgery, SVS, AVF, CT Innovation, VSGNE, NIH, Boston Scientific, Medtronic, EnVVeno Medical, Inari Medical.

## Declaration of AI and AI-assisted technologies in the writing process

During the preparation of this work the author(s) used ChatGPT-5 in order to improve readability and language. After using this tool/service, the author(s) reviewed and edited the content as needed and take(s) full responsibility for the content of the publication.

## Notes

### Competing Interest Statement

The authors have declared no competing interest.

### Author Declarations

The Human Investigations Committee at Yale University gave ethical approval for this work.

### Summary of Updates

Update with multiple sensitivity analyses to further test the robustness of results.

