## Supplementary material for "Identification of multi-omic pleiotropy factors for peripheral artery disease": ST1-3 and SF1

**Supplemental Table 1 Outcome definition codes in UK Biobank.**

|  | Self-report disease | ICD9 | ICD10 | Self-report operation | OPCS | Others |
| --- | --- | --- | --- | --- | --- | --- |
| PAD | 1067, 1087, 1088 | 4400, 4402, 4438, 4439 | I70.00, I70.01,  I70.20, I70.21,  I70.80, I70.90, I73.8, I73.9 | 1102, 1108, 1440 | X09.3, X09.4, X09.5 L21.6, L51.3, L51.6, L51.8, L52.1, L52.2, L54.1, L54.4, L54.8 L59.1, L59.2, L59.3, L59.4, L59.5, L59.6, L59.7, L59.8, L60.1, L60.2,  L63.1, L63.5, L63.9, L66.7 |  |
| T2D | 1223 |  | E11 |  |  | 2443 |
| CAD | 1075 | 4109, 4119, 4129, 429.79 | I21-I23, I24.1, I25.2 | 1075, 1095 | K40, K41, K45, K49, K50.2, K75 |  |

Supplemental Table 2 Colocalization results.

| Pleiotropy pair | Variant | Gene | Number of SNPs | PP.H0.abf | PP.H1.abf | PP.H2.abf | PP.H3.abf | PP.H4.abf |
| --- | --- | --- | --- | --- | --- | --- | --- | --- |
| PAD-T2D | eQTL | C4A | 2522 | 1.83E-14 | 1.34E-12 | 1.28E-02 | **9.38E-01** | 4.92E-02 |
| PAD-T2D | eQTL | DDAH2 | 249 | 2.80E-12 | 1.36E-11 | 1.33E-02 | 6.35E-02 | **9.23E-01** |
| PAD-T2D | pQTL | PLEKHA1 | 299 | 6.59E-09 | 2.37E-10 | **9.61E-01** | 3.46E-02 | 4.91E-03 |
| PAD-T2D | pQTL | APOE | 19 | 2.93E-11 | 1.05E-12 | 4.88E-02 | 8.04E-04 | **9.50E-01** |
| PAD-CAD | eQTL | CTD-2228A4.1 | 101 | 3.28E-02 | 7.43E-04 | 3.64E-01 | 7.64E-03 | **5.95E-01** |
| PAD-CAD | eQTL | LPL | 268 | 5.25E-13 | 3.52E-12 | 1.70E-02 | 1.13E-01 | **8.70E-01** |
| PAD-CAD | pQTL | STAT3 | 560 | 1.62E-05 | 3.03E-05 | 1.90E-01 | 3.56E-01 | **4.54E-01** |
| PAD-CAD | pQTL | PLEKHA1 | 283 | 1.31E-03 | 4.79E-05 | **6.25E-01** | 2.25E-02 | 3.51E-01 |

Supplemental Table 3 PLACO+ results.

| D2 | SNP/Gene | T.placo.plus | p.placo.plus | study |
| --- | --- | --- | --- | --- |
| T2D | rs2138161 | 66.87 | 9.11E-23 | GWAS |
| T2D | rs1333045 | 96.05 | 6.30E-32 | GWAS |
| T2D | rs582118 | 34.85 | 1.18E-12 | GWAS |
| T2D | rs149027146 | 22.35 | 1.21E-08 | GWAS |
| T2D | rs59956089 | 42.36 | 4.90E-15 | GWAS |
| T2D | DDAH2 | 22.28 | 9.27E-07 | TWAS |
| T2D | C4A | 26.94 | 7.09E-08 | TWAS |
| T2D | PLEKHA1 | -12.36 | 2.05E-04 | PWAS |
| T2D | APOE | -12.27 | 2.16E-04 | PWAS |
| CAD | rs932054 | 31.21 | 1.51E-11 | GWAS |
| CAD | rs2138161 | 34.52 | 1.31E-12 | GWAS |
| CAD | rs6841581 | 71.60 | 2.05E-24 | GWAS |
| CAD | rs9296133 | 31.61 | 1.13E-11 | GWAS |
| CAD | rs144078421 | 80.76 | 2.60E-27 | GWAS |
| CAD | rs4646272 | 58.09 | 3.94E-20 | GWAS |
| CAD | rs79390162 | 99.31 | 3.57E-33 | GWAS |
| CAD | rs9295128 | 195.26 | 2.05E-63 | GWAS |
| CAD | rs386453 | 55.44 | 2.75E-19 | GWAS |
| CAD | rs143665477 | 49.01 | 3.04E-17 | GWAS |
| CAD | rs182443492 | 48.91 | 3.29E-17 | GWAS |
| CAD | rs10455872 | 422.63 | 6.73E-135 | GWAS |
| CAD | rs140570886 | 242.30 | 3.24E-78 | GWAS |
| CAD | rs6905073 | 52.75 | 1.97E-18 | GWAS |
| CAD | rs1835346 | 32.42 | 6.20E-12 | GWAS |
| CAD | rs143843429 | 101.81 | 5.79E-34 | GWAS |
| CAD | rs2107595 | 66.88 | 6.45E-23 | GWAS |
| CAD | rs3918226 | 60.73 | 5.72E-21 | GWAS |
| CAD | rs7041637 | 121.63 | 3.21E-40 | GWAS |
| CAD | rs16905599 | 35.84 | 4.92E-13 | GWAS |
| CAD | rs4977574 | 461.45 | 4.30E-147 | GWAS |
| CAD | rs11189490 | 33.28 | 3.27E-12 | GWAS |
| CAD | rs1892971 | 32.39 | 6.31E-12 | GWAS |
| CAD | rs4766578 | 76.76 | 4.78E-26 | GWAS |
| CAD | rs17696736 | 61.04 | 4.59E-21 | GWAS |
| CAD | rs55988292 | -26.82 | 3.95E-10 | GWAS |
| CAD | rs12149545 | 31.55 | 1.18E-11 | GWAS |
| CAD | rs1893250 | 30.46 | 2.64E-11 | GWAS |
| CAD | rs2110135 | 31.93 | 8.87E-12 | GWAS |
| CAD | rs111516643 | 25.11 | 1.42E-09 | GWAS |
| CAD | rs73015007 | 46.47 | 1.97E-16 | GWAS |
| CAD | rs55997232 | 87.06 | 2.63E-29 | GWAS |
| CAD | rs4804573 | 41.87 | 5.80E-15 | GWAS |
| CAD | CTD-2228A4.1 | -11.23 | 6.54E-04 | TWAS |
| CAD | LPL | 29.76 | 3.14E-08 | TWAS |
| CAD | PLEKHA1 | 15.80 | 7.20E-05 | PWAS |
| CAD | STAT3 | 15.24 | 9.66E-05 | PWAS |

**Supplemental Figure 1. QQ plots for summary statistics of PAD, T2D, and CAD.**

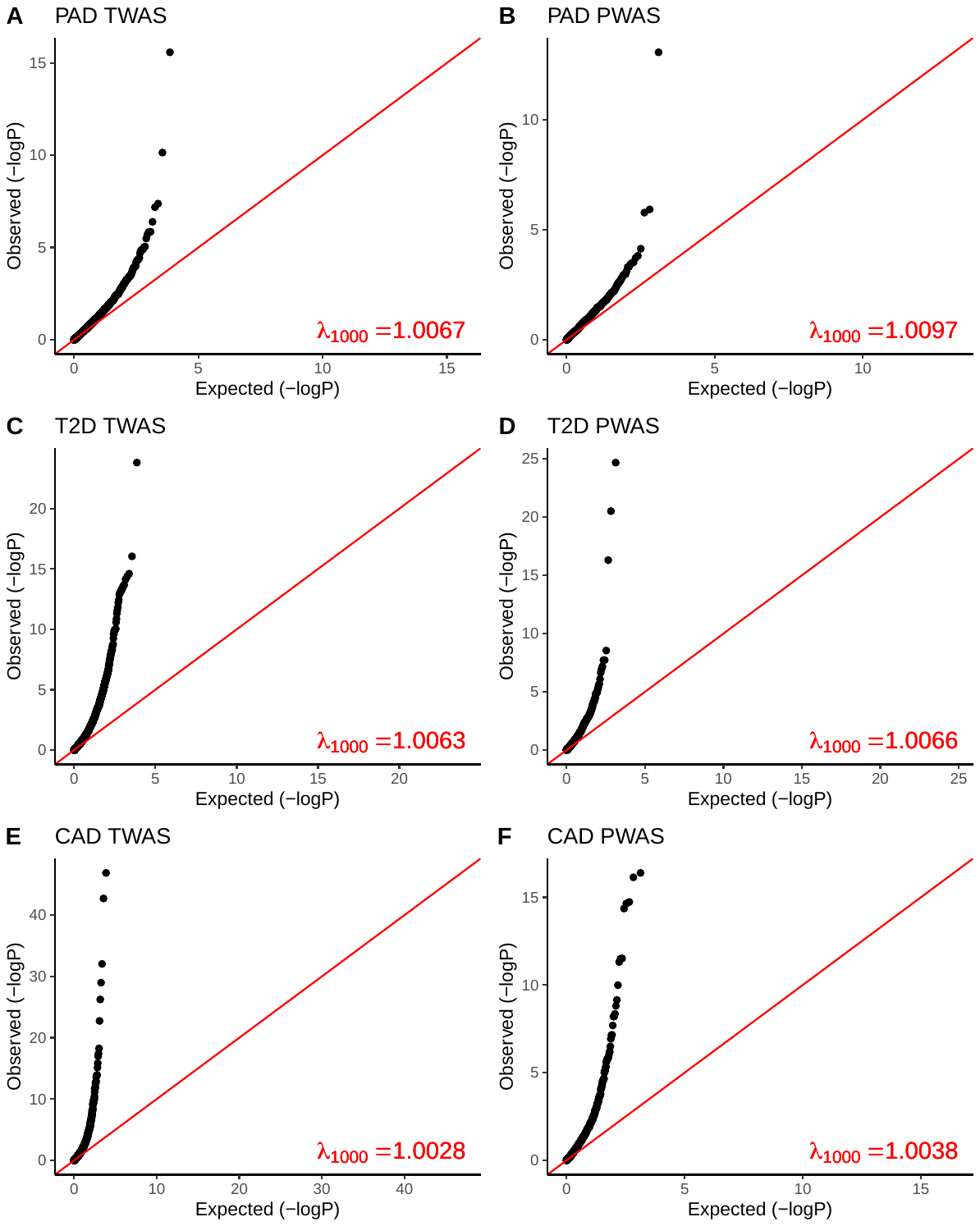

The quantile-quantile plots for GWAS, TWAS, and PWAS summary statistics of PAD, T2D, and CAD with 𝝀_1000_. All the results indicated that there was no biased due to population stratification with all estimates of 𝝀_1000_ below 1.05.
